# Understanding the impact of the Covid-19 pandemic on delivery of rehabilitation in specialist palliative care services: An analysis of the CovPall-Rehab survey data

**DOI:** 10.1101/2021.04.13.21255380

**Authors:** Joanne Bayly, Andy Bradshaw, Lucy Fettes, Muhammed Omarjee, Helena Talbot-Rice, Catherine Walshe, Katherine E Sleeman, Sabrina Bajwah, Lesley Dunleavy, Mevhibe Hocaoglu, Adejoke Oluyase, Ian Garner, Rachel L Cripps, Nancy Preston, Lorna K Fraser, Fliss EM Murtagh, Irene J Higginson, Matthew Maddocks

## Abstract

**Background:** Palliative rehabilitation involves multi-professional processes and interventions aimed at optimising patients’ symptom self-management, independence, and social participation throughout advanced illness. Rehabilitation services were highly disrupted during the Covid-19 pandemic.

**Aim:** To understand rehabilitation provision in palliative care services during the Covid-19 pandemic, identifying and reflecting on adaptative and innovative practice to inform ongoing provision.

**Design:** Cross-sectional national online survey.

**Setting/participants:** Rehabilitation leads for specialist palliative care services across hospice, hospital, or community settings, conducted from 30/07/20 to 21/09/2020.

**Findings:** 61 completed responses (England, n=55; Scotland, n=4; Wales, n=1; and Northern Ireland, n=1) most frequently from services based in hospices (56/61, 92%) providing adult rehabilitation. Most services (55/61, 90%) reported rehabilitation provision becoming remote during Covid-19 and half reported reduced caseloads. Rehabilitation teams frequently had staff members on sick-leave with suspected/confirmed Covid-19 (27/61, 44%), redeployed to other services/organisations (25/61, 41%) or furloughed (15/61, 26%). Free text responses were constructed into four themes: (i) fluctuating shared spaces; (ii) remote and digitised rehabilitation offer; (iii) capacity to provide and participate in rehabilitation; (iv) Covid-19 as a springboard for positive change. These represent how rehabilitation services contracted, reconfigured, and were redirected to more remote modes of delivery, and how this affected the capacity of clinicians and patients to participate in rehabilitation.

**Conclusion:** This study demonstrates how changes in provision of rehabilitation during the pandemic could act as a springboard for positive changes. Hybrid models of rehabilitation have the potential to expand the equity of access and reach of rehabilitation within specialist palliative care.

**Key Statements:** *What is already known about the topic?:*

- Guidelines recommend that rehabilitation targeting function, well-being, and social participation is provided by specialist palliative care services.
- Prior to Covid-19, there was variable provision of palliative rehabilitation in the UK. This variation was related to local service priorities, funding, and commissioning constraints.

*What this paper adds:*

- Over time, Covid-19 related disruptions forced services to reconfigure and adapt which caused fluctuations in the shared spaces in which health professionals, patients and family care givers met to participate in rehabilitation.
- These fluctuations resulted in the adoption of digital and remote forms of care which altered health professionals’ and patients’ capacity to participate in, and the equity of access to and reach of, rehabilitation.
- Covid-19 has acted as a springboard for learning, with many rehabilitation services hoping to move into the future by (re)gaining losses and integrating these with lessons learned during the pandemic.

*Implications for practice, theory or policy:*

- Recommendations are made to support extended reach and more equitable access to rehabilitation in palliative care services.
- We recommend mixed methods evaluations of hybrid models of in-person and online rehabilitation across palliative care settings.

## Background

Palliative care services have made essential contributions in responding to Covid-19 through engaging in advance care planning, producing guidance to manage symptoms, and caring for patients across hospital, hospice and community settings.^1-5^ These contributions have occurred in the context of a rapid increase in demand, leading to increased activity in hospital and home-based specialist palliative care teams, a shift from proactive to reactive end of life care, and wider provision of support and education for other health care professionals.^2^ These changes are likely to have impacted on provision of rehabilitation for people receiving palliative care.

Palliative rehabilitation encompasses function-focused care. It supports people towards optimal independence and participation in society throughout their disease, including during functional decline towards the end of life.^6^ It adopts a holistic and person-centered approach, comprising multi-professional assessment and mainly non-pharmacological interventions ^7, 8^ such as goal directed symptom management,^9^ physical activity and exercise,^10-12^ mindful movement ^13, 14^, and enablement in activities of daily living.^15, 16^ Rehabilitation plays a crucial role within palliative care to meet physical, psychosocial, and spiritual needs of people with advanced, progressive disease.^6, 8^

Guidelines recommend specialist palliative care services provide rehabilitation,^17, 18^ through physiotherapists and occupational therapists as core team members, and access to dietitians and speech and language therapists. However, rehabilitation in palliative care is not universally prioritised, with ad hoc and limited provision within specialist services.^19, 20^ This may have been exacerbated during the Covid-19 pandemic where rehabilitation is reported to have been the most commonly disrupted health service, often being deemed non-essential.^21^ Moreover, social distancing and isolation policies may disrupt rehabilitation provision, as many interventions are delivered in-person, involving touch, movement, groups, and interactions within the physical and social environment. Understanding how rehabilitation services were affected by and adapted to Covid-19 is needed to support the implementation of strategies to optimise the provision of this component of specialist palliative care in the future.

### Aims

To understand rehabilitation provision in palliative care services during the Covid-19 pandemic, identifying and reflecting on adaptative and innovative practice changes to inform ongoing provision.

## Methods

### Design

A cross-sectional national online survey grounded in an interpretive paradigm. This survey is part of the CovPall study ^2, 3, 5^ and is reported according to the STROBE^*^ and CHERRIES^*^ statements. Research ethics committee approval was obtained from King’s College London Research Ethics Committee (21/04/2020, Reference; LRS 19/20-18541 ISRCTN16561225). Completion of the survey indicated consent.

### Participants and Setting

Rehabilitation or therapy leads for specialist palliative care services providing rehabilitation across inpatient and out-patient palliative care in hospital or hospice, home palliative care, and nursing home settings in the UK.

### Sampling and recruitment

The invitation to participate was disseminated via allied health professions and palliative care key stakeholder organisations (Hospice UK Covid-19 (Clinical) Network; Sue Ryder; The Association of Chartered Physiotherapists in Oncology and Palliative Care; Palliative Rehabilitation Facebook Group) and via social media. Eligible service leads were provided with the link to the online survey.

### Data Collection

REDCap was used to build and host the survey. Data were collected through closed and free text responses (see Supplementary files 1 and 2 for full survey and procedures). The responses provided were reflections made by rehabilitation or service leads within the service/organisation in which they worked and was open between 30/07/20-21/09/2020.

### Data analysis

Descriptive analysis using SPSS (v24) was conducted to provide contextual data to inform qualitative analysis. Free text responses were analysed in NVivo (v12) using reflexive thematic analysis.^22, 23^ JB and AB familiarised themselves with the free-text data, coding inductively at a sematic level. Codes sharing similar meaning patterns were combined as categories, then similar categories were combined as themes. At this point, we recognised that our understandings of how Covid-19 impacted on rehabilitation services were resonant with the embodied-enactive clinical reasoning in physical therapy model.^24, 25^ This proposes that, in rehabilitation encounters, the lived experiences, backgrounds, expectations, and expertise of patients and professionals coalesce and are embodied and enacted in ‘intersubjective spaces’ (hereafter described as ‘shared spaces’). Guided by this model, we revisited and reflected on the data interpretively at a latent level, organising codes into higher order themes and subthemes. Finally, central organising concepts underpinning these themes were named and overarching themes were agreed. Throughout this process, JB, AB, and MM and wider members of the CovPall team acted as ‘critical friends’ ^26^ by challenging, questioning, and contributing to the interpretation of findings. Further analysis and engagement with the data occurred throughout the writing process. We adopted a relativist approach to rigour, selecting quality criteria applicable to the study aims and methodology^27^ (Table 1).

**Table 1.**
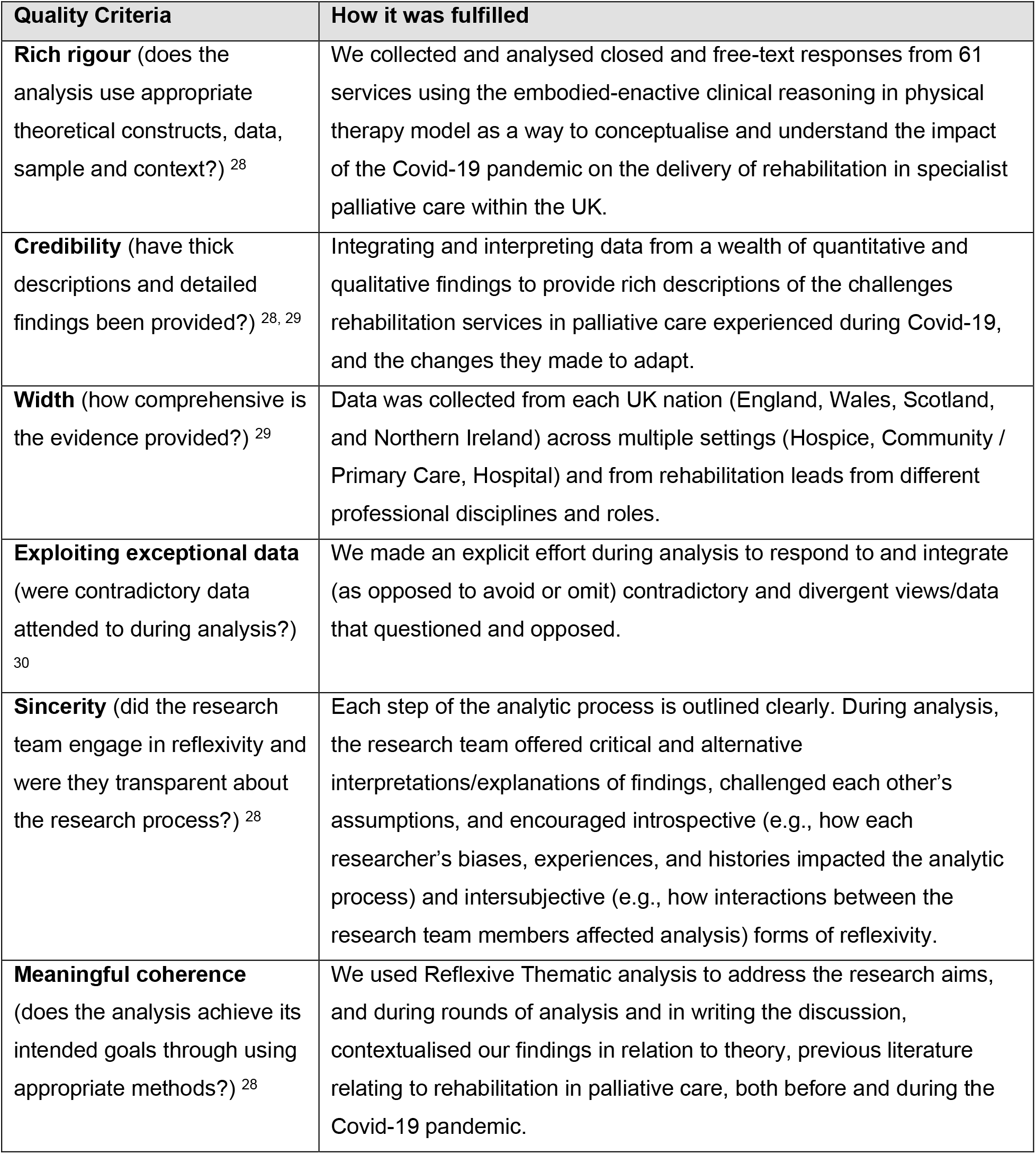
Quality criteria used and applied to ensure rigour during qualitative analysis.

## Findings

### Characteristics of services and respondents

61 completed responses were received. Characteristics of services described are presented in Table 1. Closed text responses:

Services were most frequently based in hospices (57/61), which in the UK are usually physical buildings in the charitable sector. Other services were based in the community or hospital. Staffing establishments were small; full time equivalents for physiotherapists were slightly higher than occupational therapists. Dietitians and speech and language therapists were accessed through external providers. Prior to Coivd-19 more than three-quarters of services provided rehabilitation in hospice day therapy, hospice inpatient, and hospice outpatient settings. About two-thirds provided rehabilitation in peoples’ homes, and one-third in nursing/residential care homes. A large reduction in rehabilitation provision in hospice day therapy and outpatient settings occurred. Sixteen (27%) fewer services provided rehabilitation to hospice inpatients and only 3 services continued rehabilitation provision to nursing/residential care homes (Table 2).

**Table 2.** Characteristics of services respondents.

|  |  | n/N | % |
| --- | --- | --- | --- |
| Country | England | 55/61 | 90 |
|  | Scotland | 4/61 | 7 |
|  | Wales | 1/61 | 2 |
|  | Northern Ireland | 1/61 | 2 |
| Organisation service based in | Hospice | 57/61 | 93 |
|  | Community / Primary Care | 2/61 | 3 |
|  | Hospital | 2/61 | 3 |
| Team lead professional discipline | Allied Health Professional | 41/61 | 67 |
|  | Nurse | 15/61 | 25 |
|  | Social worker | 1/61 | 2 |
|  | Physician/ medical doctor | 1/61 | 2 |
|  | Clinical director/head of service | 3/61 | 5 |
| Rehabilitation team staffing including externally contracted providers (full time equivalent, (FTE, median, range) | Physiotherapists | 1.6 (0-3.6) |  |
|  | Occupational Therapists | 1.2 (0-4.7) |  |
|  | Therapy Assistants | 0.8 (0.8-6.6) |  |
|  | Dietitians | 0 (0-3) |  |
|  | Speech and Language Therapists | 0 (0-1) |  |
|  | Other (volunteers, administrative) | 0 (0-4.4) |  |
|  | All staff members | 4.2 (0-12.3) |  |
| Age of patients cared for | Adults only | 56/61 | 92 |
|  | Adults and children | 5/61 | 8 |
| Conditions rehabilitation provided for* | Advanced (any) | 53/61 | 87 |
|  | Respiratory | 42/61 | 69 |
|  | Cancer | 44/61 | 72 |
|  | Cardiovascular | 40/61 | 66 |
|  | Neurological | 42/61 | 69 |
|  | Renal / liver | 40/61 | 66 |
|  | Dementia | 32/61 | 52 |
|  | Severely ill or dying from Covid-19 alone | 10/61 | 6 |
|  | Pre-existing condition and Covid-19 | 29/61 | 48 |
|  | Recovering from Covid-19 | 25/61 | 41 |
| Usual settings for rehabilitation prior to Covid-19* | Patient's home | 37/61 | 61 |
|  | Nursing / residential care home | 18/61 | 30 |
|  | Community hospital | 2/61 | 3 |
|  | Non-health community centre | 4/61 | 7 |
|  | Primary Care centre | 3/61 | 5 |
|  | Hospital In- / outpatients | 4/61 | 7 |
|  | Hospice In-patients | 50/61 | 82 |
|  | Hospice outpatients | 46/61 | 75 |
|  | Hospice day therapy | 51/61 | 84 |
| Settings for rehabilitation for people with suspected, confirmed or recovering from Covid-19 | Patient's Home | 21/61 | 34 |
|  | Nursing / residential care home | 3/61 | 5 |
|  | Community hospital | 1/61 | 2 |
|  | Non-health community centre | 0/61 | 0 |
|  | Primary Care centre | 1/61 | 2 |
|  | Hospital In- / outpatients | 2/61 | 3 |
|  | Hospice inpatients | 34/61 | 56 |
|  | Hospice outpatients | 3/61 | 5 |
|  | Hospice day therapy | 1/61 | 2 |

\*Respondents were able to select more than one option

Most services (55/61,90%) reported the Covid-19 pandemic had changed rehabilitation provision. Rehabilitation teams had staff members on sick leave with suspected or confirmed Covid-19 (27/61,44%), redeployed to other services/organisations (25/61,41%) or furloughed (15/61,26%). Other challenges included having difficulties providing rehabilitation equipment (23/61,38%), problems accessing personal protective equipment (PPE) (18/61,30%), and having no access and/or training in remote technologies (8/61,14%). Half of responding services reported a reduced number of referrals and caseload, and almost all reported a large shift from face-to-face to remote contacts (Figure 1).

**Figure 1.**
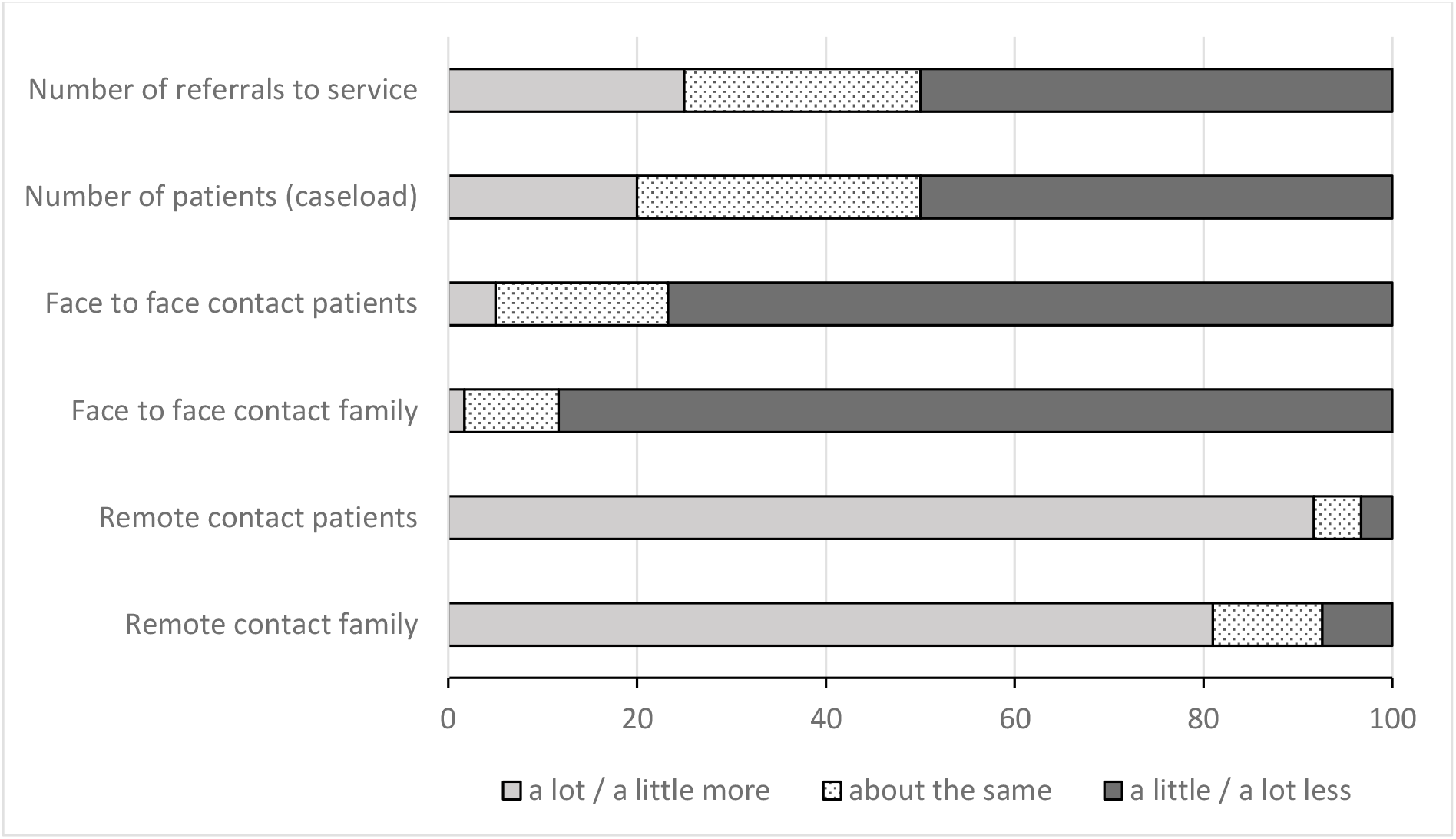
Changes to rehabilitation provision in specialist palliative care services during the Covid-19 pandemic.

Free text responses:

The analysis of free-text responses is represented by four themes and three sub-themes which outline respondents’ perceptions of the impact Covid-19 had on the organisation, delivery, and provision of palliative rehabilitation. They are represented in accordance with the embodied-enactive clinical reasoning in physical therapy model^24^ (Figure 2).

**Figure 2:**
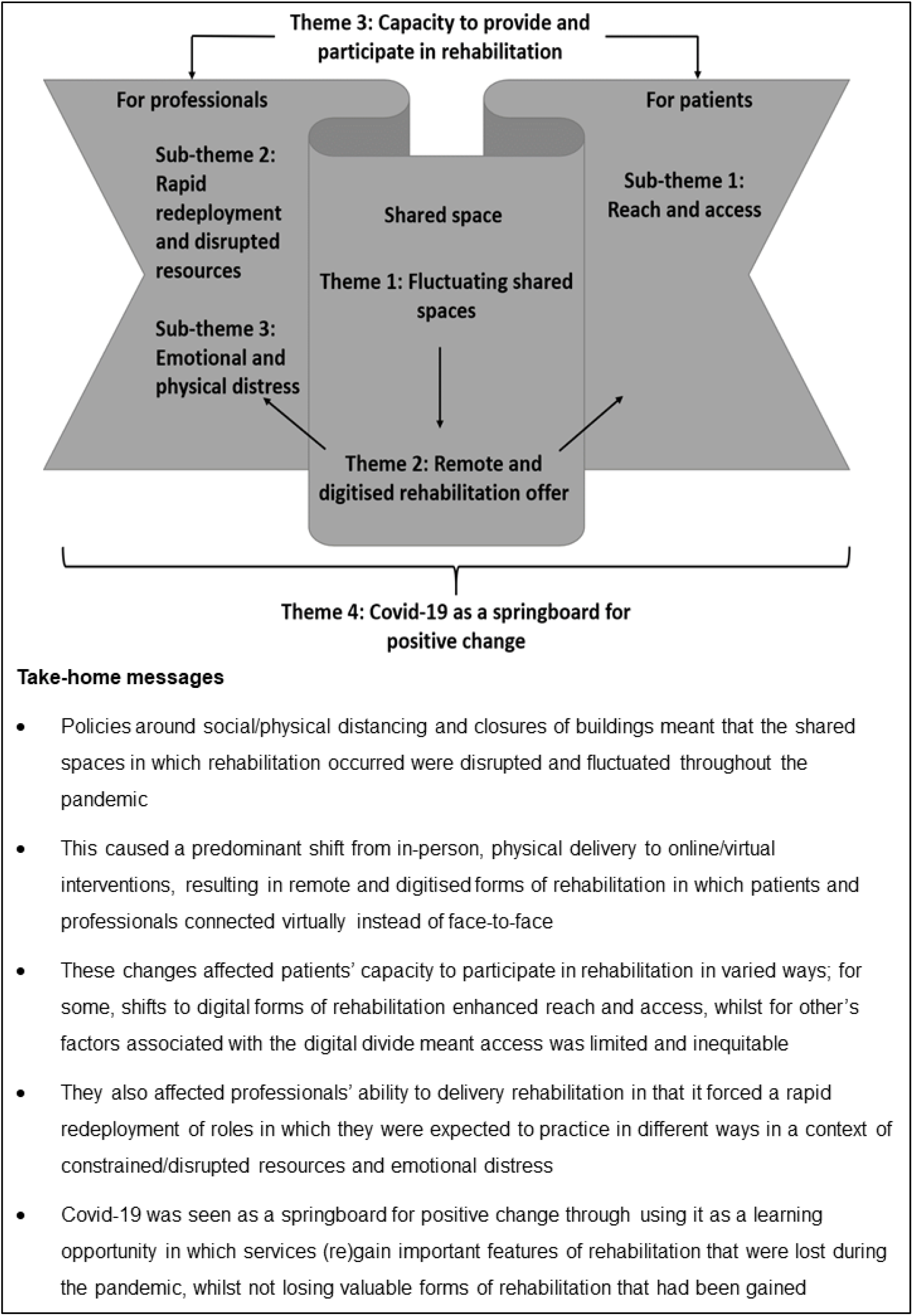
An overview of findings from free-text responses in accordance with the adapted embodied-enactive clinical reasoning in physical therapy mode.

#### Theme 1: Fluctuating shared spaces

The pandemic forced a shift in the shared spaces in which healthcare professionals and patients met to participate in palliative rehabilitation. This was due to the dangers associated with the spreading of Covid-19, thus the social and physical distancing needed to limit transmission and minimise risk. Initially, the main shift observed was away from face-to-face shared spaces to an online, virtual space as buildings were closed, staff were shielded or furloughed, and home visits stopped.

> *“Adoption of telephone and videocall assessment and intervention, no outpatient appointments or community visits offered for first 5 months of pandemic” (ID59 England)*
>
> *“Reduction in OT support in own homes (with reduced staffing + shielding). Sadly, in the early weeks, a few patients with COVID-19 and severe symptoms were unable to have Physiotherapy due to lack of appropriate PPE” (ID19, England)*

Whilst for most, shared spaces shifted to virtual platforms, a small number of services reported minimal or no changes to service provision from the outset of the pandemic. Instead, they commented on how health professionals continued to provide routine rehabilitation services in patients’ homes and inpatient units:

> *“No change in practice however we have continued to give see and treat patients using all the correct guidelines and PPE if the patient consents. Therefore, we have been able to give a continuous service unlike our community colleagues who have been restricted in their service” (ID15 England)*

Shifts in the shared spaces were dynamic and fluctuated throughout the pandemic as more was learned about Covid-19, government alert levels were altered, and resources and risk assessment systems were developed. Some services that had initially moved all rehabilitation to virtual spaces began to reintroduce limited in-person rehabilitation in community and inpatient settings when deemed ‘essential’ following risk assessments and dependent on the availability of PPE.

> *“As Alert level decreased allowed to see patients in their own home with appropriate risk assessment and PPE” (ID08 England)*
>
> *“Initially no face to face on the in-patient unit but now that has resumed on a reduced basis” (ID03 England)*

#### Theme 2: Remote and digitised rehabilitation offer

Integral to palliative rehabilitation was providing interventions in-person where embodied interactions (e.g., touch, movement, group-based activities) with people and the surrounding physical/social environment were fundamental. Fluctuations in shared spaces led to changes in the ‘rehabilitation offer’ (i.e., what and how interventions were delivered). Respondents highlighted how closures of buildings and physical spaces resulted in shifts to long-range forms of rehabilitation. This represented how the physical, embodied and enacted in-person components through which rehabilitation was usually delivered was replaced by video-conferencing platforms in which patients and professionals connected, and interventions, assessments, and group therapies were delivered, digitally.

Delivering rehabilitation through digital means required services to adapt creatively by thinking of different ways they could support people with common symptoms and concerns (e.g., breathlessness, anxiety, and fatigue). These adaptations were not uniform; some interventions adopted a synchronous approach (e.g., providing live group-based classes via Zoom), whilst others were asynchronous (e.g., uploading previously or newly produced patient facing resources to websites or YouTube).

> *“use of AccRx on SystmOne* [clinical online virtual platforms] *for video consultations, sending out more postal information to patients. Zoom recorded and live groups sessions” (ID09 England)*
>
> *“We have been developing online versions of our groups such as Tai Chi and Fatigue and Breathlessness, these have started running recently. Advice and exercises have been posted out to individuals and we have also used accuRx for 1:1 video assessment and treatment as indicated” (ID30 England)*
>
> *“The wellbeing service has become a virtual service providing support calls/video consultations. Support groups for patients through the use of zoom such as the Be in Charge programme which provides tailored support for anxiety management/ fatigue / breathlessness etc. This involves members of the multi-disciplinary team. Lymphoedema services completing initial patient assessment via telephone/video consultation prior to face to face for hosiery measuring” (ID22, England)*

Moreover, responding to Covid-19 also entailed being creative in engaging family members in the rehabilitation process (i.e., through supporting occupational therapy assessments in patients’ homes):

> *“All community visits reduced and photos, relatives measuring furniture used as first line instead” (ID45, England)*

#### Theme 3: Capacity to provide and participate in rehabilitation

Fluctuations in shared spaces and shifts to predominantly remote offers of rehabilitation had consequences for the capacity for health professionals to provide, and patients to participate in, rehabilitation.

#### For patients

##### Sub-theme 1: Reach and access

Respondents provided varied accounts on the impact that moving towards remote/digital forms of rehabilitation had on the capacity for patients to engage in rehabilitation during the pandemic. Some respondents perceived that this new way of working enhanced access, meaning that rehabilitation teams could expand their reach to people they had not been able to reach before (e.g., people in rural areas, younger people, or those too ill/unable/unwilling to travel to the hospice building):

> *“some have said that the effort of transferring to a car and then visiting the building can be very demanding on them and virtual input has proved more efficient for them. Family members have also not needed to find someone to sit with the person they care for” (ID30 England)*
>
> *“The changes have largely enabled a very small palliative rehabilitation team to expand their reach” (ID13, England)*

However, changes were not always equitable. Concerns were raised that a digital divide limited the capacity of many patients to participate in rehabilitation, especially those with communication/cognitive difficulties or with no access to computers/internet. Others lacked the ability to navigate these platforms or did not like digital forms of care delivery:

> *“Physical access has been reduced and transport has not been provided or restricted. Some patients don’t have the ability to access technology in order to have online appointments” (ID07, England)*
>
> *“Those with communication and or cognitive difficulties especially if don’t have access to video technology or lack or other to advocate for them are finding access hard and communication when wearing masks difficult” (ID61, England)*

There were also concerns that the reach of digital forms of rehabilitation were somewhat limited because certain interventions required clinicians to be physically present and use sensory cues to assess patients in ways that were not possible virtually. Respondents also voiced apprehension about how the lack of face-to-face services combined with a limited availability of PPE meant some patients in the community could not always be seen and missed out on important rehabilitation input.:

> *“Sadly in the early weeks, a few patients with COVID-19 and severe symptoms were unable to have Physiotherapy due to lack of appropriate PPE” (ID19, England)*
>
> *“Specific treatments can only be offered if seen visually otherwise general advice will be given” (ID29, England)*
>
> *“It feels as though there are a lot of patients out there in the community who are slipping through the net at present. We know they are out there but due to shielding and changes to general community input we are struggling to find patients not already known to the Hospice/service” (ID08, England)*

#### For healthcare professionals

##### Sub-theme 2: Rapid redeployment and disrupted resources

Participants reported that, in responding to fluctuations in shared spaces, various forms of rapid redeployment occurred. As the buildings/places in which they usually provided rehabilitation were closed, rehabilitation staff were redeployed to support wider members of the multi-disciplinary team. In some cases, staff used this as an opportunity to promote and provide rehabilitative approaches in other contexts (e.g., online and in-patient units). In others it included providing input where other community services had been withdrawn.

> *“All AHP/Rehab staff furloughed. Redeployed to NHS” (ID47 Hospice, Scotland)*
>
> *“Other community services locally no longer supporting/working in the way they usually would and therefore workload has increased in supporting complex needs at home. Hospice at Home service has increased and therefore required increased support from physiotherapy and occupational therapy” (ID05 England)*
>
> *“During the peak of the outbreak at the hospice, OT’s and physios supported the provision of essential care at the hospice, working bank holidays as health care assistants or managing incoming telephone calls with family members” (ID37 Wales*)

These forms of redeployment affected health professionals’ capacity to provide rehabilitation in numerous ways. For some, this was attributable to varied degrees of self-confidence that staff possessed in developing new models of rehabilitation with little time to train or adapt. There were also concerns about the practicalities involved in supporting patients to use the technologies as well as data protection and security:

> *“Finding the optimum way of using it and the practicalities of demonstrating exercises on screen” (ID03 England)*
>
> *“Lack of understanding of GDPR* [General Data Protection Regulations] *for which ones we can use, lack of access to technology for both patients and staff, unable to go to patients to teach them how to use technology (particularly at the start of the pandemic”) (ID07 England)*

Respondents voiced concerns that rapid redeployment of roles and practices undermined their perceived capacity to provide effective palliative rehabilitation, particularly when delivering it digitally. This was because digital/virtual approaches omitted the hands-on care and non-verbal forms of communication that they considered as fundamental to rehabilitation. Moreover, not every service had been able to adapt interventions in a form that could be delivered remotely:

> *“It has been difficult to connect with patients via a screen if they are upset. Normal reliance on nuanced body language and tone of voice has been hampered so needs to be approached differently. In addition, telling a group that one of their members has died has been difficult without the opportunity to approach individuals differently (sometimes in face to face we may choose to take a group member aside to break the news). Not being able to offer comforting touch is difficult” (ID14, England)*
>
> *“Not having face to face does mean you lose something with the client, that therapeutic connection. Hands on assessment is missing” (ID54 England)*
>
> *“Do not yet have a wide range of videos or presentations to cover all usual aspects of a self-management programme” (ID02 England)*

Confounding the issues associated with rapid redeployment and working differently for health professionals, was operating in a context of disrupted resources. Respondents sensed that palliative rehabilitation was sometimes viewed as dispensable/non-essential, with constraints on timely access to external equipment providers undermining their capacity to source equipment that was important for patients to function independently.

> *“We do not provide equipment but normally have good relations with local teams who provide this. these teams are working differently and those with general rehab needs are not being seen as they are not at a high enough priority for their current service offering”* (ID30 England)
>
> *“Equipment services are not delivering non-essential equipment in the community. Wheelchair services now have a 9-12 month wait for a review of a patient’s seating/wheelchair”*.*” We’ve had to set up our own buffer store to address this” (ID56 Scotland)*

At times, patients were advised to avoid equipment, to go without, or the responsibility for acquiring the equipment was shifted to individual patients:

> *“Used stock from store cupboard, advised patients on strategies avoiding equipment. Some patients purchased their own online” (ID12, England)*

##### Sub-theme 3: Emotional and physical distress

Health professionals’ capacity to deliver palliative rehabilitation was also influenced by the emotional and physical impact (e.g., fear, uncertainty, anxiety, stress, exhaustion, frustration, and burnout) of working in the context of the pandemic. For some respondents, the source of emotional distress was a consequence of attempting to fulfil job roles in a context of disorientation, general uncertainty, rapid changes to ways of working, and fears over Covid-19:

> *“Anxiety within team about the virus. Uncertainty due to differing local policies i*.*e. other community teams, etc’ (ID05, England)*
>
> *“The exhaustion and disorientation felt in the early days where the situation was rapidly evolving was particularly difficult and stressful for all involved” (ID37, Wales)*

For others, emotional and physical distress was directly related to the changes in rehabilitation. Covid-19 meant that the places and spaces in which teams could operate contracted, fracturing valued in-person communication with patients, families, and team members, and disrupting integrated working between teams and services:

> *“half the team had the infection which increased team anxieties, stopped a level of patient care, delayed some patient assessments due to sickness and isolation timescales” (ID25, England)*
>
> *“Managing morale. Team feeling more isolated. Dealing with not being able to see patients face to face and deliver normal service*… *Not being in their usual workspaces. Not seeing some colleagues for months. Zoom fatigue, Covid fatigue and resilience” (ID03, England)*

Moreover, some respondents highlighted distress associated with a lack of transparency over their own and others job security. Over time, these issues disrupted capacity by leading to worsening mental health, degraded morale/motivation and, in some cases, staff leaving roles.

> *“communication from hospice to furloughed staff has been poor, frustration outpatient services is not opening any time soon, some social media comments from public about lack of rehab service has been noted physios are looking at other employment due to their treatment unsure if redundancies is a possibility” (ID60, England)*
>
> *“increased anxiety around job security and changes to the hospice. Tension in the team due to disjointed and remote working. increased workload on remaining therapists” (ID36, England)*

#### Theme 4: Covid-19 as a springboard for positive change

Responding to survey questions related to innovations and the future, respondents focused on how palliative rehabilitation services could use the pandemic as a springboard for positive change. This was through regaining aspects of rehabilitation that patients valued but were lost due to the pandemic (e.g., face-to-face interventions), whilst simultaneously not losing the valuable forms of rehabilitation that had been gained. Respondents recommended capitalising on health professionals’ newfound competencies, skills, and confidence in delivering rehabilitation remotely by developing hybrid approaches that could reach more patients and with savvy use of health professionals’ time and resources:

> *“Virtual groups, video consultations, more satellite clinics, better use of time and physical resources. It has given us time to reconsider how to deliver services to increase reach to more patients but less intensive and less site based (perhaps appropriately so)” (ID61, England)*
>
> *“We are hoping to become more integrated with day therapy services with their nurses looking at becoming more rehabilitation focussed. The senior management team has had an opportunity to look at space and there will be the development of a separate rehabilitation space with more outpatients, gym groups, videoed sessions and virtual groups” (ID13, England)*

Respondents also saw value in maintaining developments in integrated team working and collaborations that had been nurtured during the pandemic. For some, potential benefits were seen at a regional level in continuing collaborative working across hospice teams by pooling resources and skillsets in order to provide more comprehensive rehabilitative services. For others, value was seen in maintaining more local collaborations to complement rehabilitation services, including drawing on community groups to support rehabilitation in the community, upskilling volunteers, and involving the multi-disciplinary hospice team in rehabilitation conversations/interventions:

> *“Closer MDT working now. We’re starting to do more assessments with nurses to see people earlier rather than waiting for referral. Physio will be leading on the respite and rehab service from mid-October”* (ID07, England)
>
> *“The focus over the last few months has been in maintaining essential community services for patients amidst concerns about systems being overwhelmed and staffing levels being depleted. This has meant a reorganising of services to a regional rather than hospice level with collaboration of community teams across several hospices. The focus of this has not been on rehabilitation - possibly as other hospices have a less developed rehabilitation service and possibly because of concerns about resources during the pandemic. The result has been the development of a reactive rather than proactive service with no focus on rehabilitation. However, in the long term, the potential benefits of this collaborative working may be in having the ability to provide more comprehensive rehabilitation services across several hospices by pooling resources and this is something I hope to start discussing very soon” (ID17, England)*

### Discussion

#### Main findings/results of the study

This study demonstrated how Covid-19 disrupted the shared spaces in which rehabilitation in specialist palliative care was conducted. The shutting of buildings and physical spaces in which rehabilitation usually took place, combined with policies around physical/social distancing, predominantly resulted in the adoption of remote and digitised rehabilitation processes. This had mixed impacts on the capacity of health professionals to deliver, and patients’ ability to participate in, rehabilitation. Despite the disruptions and challenges that Covid-19 caused, many respondents reflected on how the pandemic could act as a springboard for positive future change through the adoption of hybrid rehabilitation approaches and the continuation of integrated /collaborative working.

#### What this study adds

This is the first study to collect empirical data that shows how the Covid-19 pandemic impacted the provision of rehabilitation in specialist palliative care services, alongside identifying innovative practice changes to inform future provision. It builds on previous work by the CovPall team ^2, 3, 5, 31^ in developing a comprehensive picture of how palliative care services responded to the Covid-19 pandemic and contributes to the literature in three ways.

First, the Covid-19 pandemic severely disrupted rehabilitation services within palliative care. The shared spaces in which rehabilitation usually took place were no longer viable, and workforce capacity was limited by staff shielding, sickness, and redeployment. Rehabilitation services contracted, reconfigured, and redirected. These findings expose the vulnerability of clinical teams providing rehabilitation in palliative care services. Teams are usually small in number and were already operating in a national context of underinvestment^32^ which left little slack in the system to deal with the rapid demands imposed by Covid-19. Operating in understaffed and under-resourced services meant the capacity to provide rehabilitation was limited. This resonates with the World Health Organisation global rapid assessment of service provision for non-communicable diseases which found that rehabilitation was the most commonly disrupted healthcare service during the Covid-19 pandemic^21^. Within this disrupted context, respondents sensed that rehabilitation services were perceived as non-essential. That respondents felt the provision of rehabilitation was under prioritised and resourced during the pandemic is concerning. Increased demand is expected to continue as Covid-19 resulted in people presenting late with advanced symptomatic disease, compounded by shielding related deconditioning,^33^ cancellations/delays in treatments and long-Covid.^34-36^ The value of rehabilitation as part of palliative care’s holistic approach should be recognised, implemented, and resourced accordingly.

Second, this work underscores the inequities regarding the ability of patients in rural/remote geographic areas, or those who are too ill to travel, to access on-site rehabilitation services in palliative care.^37, 38^ This highlights how the Covid-19 pandemic compounded already-existing inequities in palliative care^39^and contributes novel insight into the ways in which it shifted inequities. That is, as rehabilitation provision moved to virtual platforms, for people who had previously struggled to attend in-person appointments and had access to/skills to use digital technologies, access to rehabilitation improved. In contrast, for those without access to/skills to use digital technologies, and/or were shielding and unable/unwilling to risk in-person appointments, access worsened. These align with previous work in palliative that has demonstrated how shifts to online/digital service delivery has the potential to improve access for some, but worsen it for others,^40, 41^ and exemplify how the digital divide has led to new inequities in the provision of palliative rehabilitation as services moved to remote forms of provision to compensate for the Covid-19 pandemic.^42, 43^

Third, our findings highlight ways in which people working in rehabilitative palliative care services felt that Covid-19 could act as a springboard for positive future change. Covid-19 created a ‘forced shift’ to virtual working in which services and staff developed a digital confidence that, in some instances, enabled them to meet increasing demand.^44^ Indeed, the pandemic seemed to present numerous ‘teachable moments’ ^45^ in which, despite considerable challenges, respondents recognised the potential of harnessing learning through the adoption of hybrid approaches (e.g., blended face-to-face and remote provision) in future care. Digital models of care that extend reach and meet increased demand are promising ventures in reshaping and re-envisioning future rehabilitation towards more sustainable forms of palliative care. However, it is important that research and community engagement underpin these shifts to ensure that hybrid models are developed and delivered in equitable, culturally congruent, and person-centred ways that do not perpetuate already existing, or create new forms of, inequities in palliative care^39^. Studies should build on evidence for remote rehabilitation in cancer ^46, 47^ and chronic respiratory disease^48^, with robust and theoretically informed studies of digital health interventions in palliative care.^49, 50^ The pandemic provides an opportunity for palliative care services to reflect on the provision of care directed to optimising function^20^. Rehabilitation should not be limited to the therapies allied health professionals provide, it is a process requiring integrated multi-professional teams with rehabilitation expertise ^51^ as exemplified by holistic breathlessness services.^9^

#### Strengths and limitations of the study

This paper has several strengths. With responses from rehabilitation leads at 61 palliative care services, the findings represent the practice of hundreds of clinicians involved in the provision of palliative rehabilitation and the breadth of responses is large. Our methodology was robust. Researchers, palliative care clinicians and members of the public contributed to the survey development and refinement of survey questions following the first CovPall Survey. Two researchers, with contributions from the wider CovPall team, used robust and rigorous qualitative methods underpinned by theory. A balance was achieved between closed and open responses in the survey and analysis, with space provided for people to report rich data. Regarding potential limitations, it is possible the survey did not capture views of all rehabilitation team members, as it was completed by team leads. Most responses came from hospices and it is not clear if this reflects non-responses or the absence of palliative rehabilitation from other palliative care settings. We cannot ascertain from our data how our findings varied across organisations according to local contractual arrangements for the provision of rehabilitation.

### Conclusion

This study provides evidence of the impact that Covid-19 had on rehabilitation services working in palliative care within the UK. The pandemic forced shifts to remote provision and impacted the capacity of health professionals and patients to deliver and participate in rehabilitation. Evidence is provided on how the pandemic may act as a springboard for positive future changes through the adoption of hybrid approaches to rehabilitation that integrate remote and face-to-face provision in ways that are able to expand reach and improve equity. Empirical views of patients on the changes introduced have yet to be obtained and patients voices should inform future research around hybrid models of rehabilitation.

## Data Availability

Data sharing: Applications for use of the survey data can be made for up to 10 years, and will be considered on a case by case basis on receipt of a methodological sound proposal to achieve aims in line with the original protocol. The study protocol is available on request. All requests for data access should be addressed to the Chief Investigator via the details on the CovPall website (https://www.kcl.ac.uk/cicelysaunders/research/evaluating/covpall-study, and palliativecare{at}kcl.ac.uk) and will be reviewed by the Study Steering Group.

## Authorship

IJH is the grant holder and chief investigator; KES, MM, FEM, CW, NP, LKF, SB, MBH and AO are co-applicants for funding. IJH and CW with critical input from all authors wrote the protocol for the CovPall study. JB, MM, LF and HTR amended the CovPall Survey to produce the Covpall-Rehab survey. JB co-ordinated data collection and distributed survey with input from MM and LF. JB, AB, MM and MO analysed the data. All authors discussed the interpretation of findings and take responsibility for data integrity and analysis. JB and AB drafted the manuscript. All authors provided critical revision of the manuscript for important intellectual content. IJH is the guarantor.

## Data sharing

Applications for use of the survey data can be made for up to 10 years and will be considered on a case-by-case basis on receipt of a methodological sound proposal to achieve aims in line with the original protocol. The study protocol is available on request. All requests for data access should be addressed to the Chief Investigator via the details on the CovPall website (https://www.kcl.ac.uk/cicelysaunders/research/evaluating/covpall-study, and) and will be reviewed by the Study Steering Group.

## Acknowledgements

This study was part of CovPall, a multi-national study, supported by the Medical Research Council, National Institute for Health Research Applied Research Collaboration South London and Cicely Saunders International. We thank all collaborators and advisors. We thank all participants, partners, PPI members and our Study Steering Group. We gratefully acknowledge technical assistance from the Precision Health Informatics Data Lab group (https://phidatalab.org) at National Institute for Health Research (NIHR) Biomedical Research Centre at South London and Maudsley NHS Foundation Trust and King’s College London for the use of REDCap for data capture.

We thank all services for responding. The following indicated they were happy to be acknowledged: Arthur Rank Hospice Charity, Dorothy House Hospice Day Service, East Cheshire Hospice, Garden House Hospice care, Isabel Hospice, John Taylor Hospice, Kilbryde Hospice, Longfield Community Hospice, Marie Curie Hospice Cardiff and the Vale, Marie Curie Hospice West Midlands, Martlets Rehabilitation Team, Mary Ann Evans Hospice, Mountbatten Isle of Wight and Hampshire, Overgate Hospice, Peace Hospice Care, Pilgrims Hospices, Phyllis Tuckwell Hospice Care, Princess Alice Hospice, Rosemary Foundation, Salisbury Hospice, St Andrew’s Hospice, St Ann’s Hospice, St Catherine’s Scarborough, Southern Area Hospice Services Newry, St Andrew’s Hospice, St Barnabas House Hospices, St Christopher’s Hospice, St Cuthbert’s Hospice, St Elizabeth Hospice, St Joseph’s Hospice, St Luke’s Hospice, Sy Margaret’s Hospice, St Mary’s Hospice, St Michael’s Hospice, St Nicholas Hospice, St Wilfrid’s Hospice Eastbourne, Strathcarron Hospice, Sue Ryder South Oxfordshire Palliative Care Therapy Team, Sue Ryder Wheatfields Hospice, Sue Ryder, Leckhampton Court Hospice, Teesside Hospice Care Foundation., Wigan and Leigh Hospice.

