## Supplementary File 1 for "Understanding the impact of the Covid-19 pandemic on delivery of rehabilitation in specialist palliative care services: An analysis of the CovPall-Rehab survey data"

### COVPALL-Rehabilitation Collaboration Survey

Improving rehabilitation in palliative care services for people with and without COVID-19 by shared learning.

Thank you for considering completing this survey. We are trying to find out how rehabilitation teams within palliative care services are changing because of the COVID-19 pandemic. This is important as the disease is new and palliative care rehabilitation teams are changing how they work, so there is an opportunity to learn from each other.

We realise you are very busy right now and so we have tried to balance collecting information that patients, services and policy makers think is most helpful, while keeping the questionnaire as short as we can.

The questionnaire has 5 sections and should take no longer than 30 minutes to complete, although it may depend on how many additional/open comments you wish to share. We will consider everything you write. Although the grouped results will be shared, we will not name your rehabilitation team unless you ask us to.

You can pause the questionnaire by the clicking ‘Save and Return Later’ button at the bottom of the page. You will be given a code and sent a link to enable you to continue later. (If you forget the link, there should be a ‘returning’ option at the top right of this page.) If you wish to correct errors after you have clicked ‘Submit’, please with COVPALL-REHAB in the subject line.

If you have any concerns about this questionnaire or this study, please.
COV-PALL-REHAB is led by Professor Irene Higginson and Dr Matthew Maddocks of the Cicely Saunders Institute, with a multi-professional team of partners from different organisations and backgrounds. Patient representatives have contributed to this questionnaire and our plans. For more information see https://www.kcl.ac.uk/cicelysaunders/research/evaluating/covpall-study

This study has been granted ethical approval by the PNM Research Ethics Subcommittee of King’s College London, code LRS-19/20-18541.

For the purposes of this questionnaire, rehabilitation includes all the usual assessments and interventions provided by Allied Health Professionals and associated colleagues, including dietitians, occupational therapists, physiotherapists, speech and language therapists and therapy assistants. Interventions provided by other disciplines can be included if they are explicitly provided within a rehabilitation service.

#### 1. ABOUT YOU AND YOUR SERVICE

1.1 Contact email of the person completing the survey

__________________________________
(We need this information so we can help you get back into the survey, or help you to complete it, if you have difficulty)

1.2 Name of the person completing the survey

__________________________________

1.3 Date

__________________________________
(DD-MM-YYYY)

| | |
|---|---|
| 1.4 Country | ◯ England<br>◯ Scotland<br>◯ Wales<br>◯ N Ireland<br>◯ Australia<br>◯ Belgium<br>◯ Canada<br>◯ Germany<br>◯ Ireland<br>◯ Italy<br>◯ Poland<br>◯ New Zealand<br>◯ Other (a box will open)<br>(A regions option may appear) |
| 1.4a Country (please specify) | __________________________________ |
| 1.4b English Regions | ◯ North East<br>◯ North West<br>◯ Yorkshire and The Humber<br>◯ East Midlands<br>◯ West Midlands<br>◯ East<br>◯ London<br>◯ South East<br>◯ South West |
| 1.4c Welsh region | ◯ N Wales<br>◯ W Wales<br>◯ SE Wales<br>◯ Mid Wales |
| 1.4d Scottish region | ◯ Fife, Lothian, Borders, Dumfries & Galloway<br>◯ Greater Glasgow & Clyde, Ayrshire & Arran, Lanarkshire, Forth Valley<br>◯ Tayside, Grampian, Western Isles, Highland, Orkney and Shetland |
| 1.4e Region | __________________________________ |
| 1.5 Please specify your therapy manager role | __________________________________ |
| 1.6 Professional Discipline | ◯ Physiotherapist / Physical Therapist<br>◯ Occupational Therapist<br>◯ Dietitian<br>◯ Speech and Language Therapist / Speech Pathologist<br>◯ Nurse<br>◯ Social Worker<br>◯ Physician/medical doctor<br>◯ Non-clinical manager<br>◯ Other (a box will open) |
| Other Professional Discipline | __________________________________ |

Next Page takes you to the main questions. You must complete the information above first, but you can return here later if you wish to change anything
You must click "Next Page" or "Save and Return Later" to save any data you have entered

#### MAIN SURVEY

2. ABOUT THE REHABILITATION USUALLY OFFERED BY YOUR SERVICE BEFORE THE COVID-19 PANDEMIC

2.1 Which type of organisation is your rehabilitation service based in

- ☐ Community /primary care
- ☐ Hospital
- ☐ Hospice organisation
- ☐ Other (a box will open)

(Please tick all that apply)

Other Sector

__________________________________

2.2 Approximate number of new patients seen annually

__________________________________

(Must be a number)

2.3 Types of patients cared for

- ◯ Adults
- ◯ Children (< 18 years)
- ◯ Adults and children

2.4 In what settings did your service provide rehabilitation for people under the care of a palliative care team

- ☐ Patient's home
- ☐ Nursing home
- ☐ Residential care home
- ☐ Community hospital
- ☐ Primary care centre
- ☐ Non-health community centre
- ☐ Hospital in-patients
- ☐ Hospital out-patients
- ☐ Hospice in-patients
- ☐ Hospice out-patients
- ☐ Hospice day-therapy
- ☐ Virtual (i.e. telephone, video-link)
- ☐ Other (a box will open)
- ☐

(Please tick all that apply)

Other Settings

__________________________________

2.5 Please indicate if your service provides rehabilitation for people with the following health conditions

- ☐ Any advanced health condition
- ☐ Respiratory conditions
- ☐ Cancer
- ☐ Cardio-vascular conditions
- ☐ Neurological conditions
- ☐ Renal / Liver disease
- ☐ Dementia
- ☐ Other (a box will open)

(Please tick all that apply)

Other Health Conditions

__________________________________

Additional information about usual rehabilitation services offered before the COVID Pandemic

2.6 Rehabilitation team members (a box will open for full time equivalents)

2.6a Number of Physiotherapists

__________________________________
(A box will open for FTE)

Full time equivalent Physiotherapists

__________________________________

2.6b Number of Occupational Therapists

__________________________________
(A box will open for FTE)

Full time equivalent Occupational Therapists

__________________________________

2.6c Number of Dietitians

__________________________________
(A box will open for FTE)

Full time equivalent Dietitians

__________________________________

2.6d Number of Speech and Language Therapists

__________________________________
(A box will open for FTE)

Full time equivalent Speech and Language Therapists

__________________________________

2.6e Number of Therapy Assistants

__________________________________
(A box will open for FTE)

Full time equivalent Therapy Assistants

__________________________________

2.6f Other (please specify with numbers)

__________________________________
(A box will open for FTE)

Full time equivalents for above

__________________________________

2.7 If volunteers are usually involved in the delivery of your service, please provide details

__________________________________________

2.8 Is there anything else you want to tell us about how your service usually operates that you think it is important for us to know

__________________________________________

3. REHABILITATION PROVISION DURING THE PANDEMIC

Rehabilitation provided for patients who are NOT DIAGNOSED OR WITHOUT SUSPECTED COVID-19 during the pandemic

| | |
|---|---|
| 3.1 Has COVID-19 changed how you provide rehabilitation for the types of patients you would usually support | ◯ Yes ◯ No<br>(The following questions may change) |
| Please give details of any changes to specific rehabilitation assessments and interventions during the pandemic that would usually be provided by your service for people not suspected of having COVID19 | |
| 3.1a Physiotherapy changes | __________________________ |
| 3.1b Occupational Therapy changes | __________________________ |
| 3.1c Dietetics changes | __________________________ |
| 3.1d Speech and Language Therapy changes | __________________________ |
| 3.1e Therapy assistants (please specify where linked with a specific discipline) | __________________________ |
| 3.1f Other team members (please specify) | __________________________ |
| 3.2 What is the shortest time in days that patients are under the care of your rehabilitation service during the COVID-19 pandemic | __________________________ |
| 3.3 What is the longest time in days that patients are under the care of your rehabilitation service during the COVID-19 pandemic | __________________________ |
| Does this time include people you are still caring for | ◯ Yes ◯ No |
| Rehabilitation provided for patients who have SUSPECTED, CONFIRMED or RECOVERING from COVID-19 during the pandemic | |
| 3.4 Has the rehabilitation service developed any new risk assessment procedures to guide how the service is provided safely for people with suspected, confirmed or recovering from COVID-19 | ◯ Yes ◯ No<br>(A box will open for details) |
| Please give details of these risk assessment procedures | ________________________________ |
| 3.5 Has your service provided rehabilitation for people with suspected, confirmed or recovering from COVID-19 | ◯ Yes ◯ No<br>(The following questions may change) |

3.5a Of the patients on your caseload with suspected, confirmed or recovering from COVID-19, would you say that they were
(Tick all that apply)

☐ Patients who are severely ill or dying due mainly to COVID-19
☐ Patients with pre-existing illness / co-morbidities as well as COVID-19 who are severely ill or dying
☐ Patients who are recovering following suspected or confirmed COVID-19
☐ Other (a box will open)

---

Please specify the other types of patients ____________________

---

3.6 Which settings were rehabilitation procedures provided in for people with suspected, confirmed or recovering from COVID-19

☐ Patient's home
☐ Nursing home
☐ Residential care home
☐ Community hospital
☐ Primary care centre
☐ Non-health community centre
☐ Hospital in-patients
☐ Hospital out-patients
☐ Hospice in-patients
☐ Hospice out-patients
☐ Hospice day-therapy
☐ Virtual (i.e. telephone, video-link)
☐ Other (a box will open)
(Tick all that apply)

---

Please specify the other settings ____________________

---

3.7 Please give details of the rehabilitation interventions provided by your service for people with suspected, confirmed or recovering from COVID-19

---

Physiotherapy ____________________

---

Occupational Therapy ____________________

---

Dietetics ____________________

---

Speech and Language Therapy ____________________

---

Therapy assistants (please specify where linked with a specific discipline) ____________________

---

Other team members (please specify) ____________________

---

3.8 Did you have protocols or guidance for the provision of rehabilitation for people with confirmed or suspected COVID-19

○ Yes ○ No

| | |
|---|---|
| 3.9 What sources of information did you use | ☐ Locally developed guidance<br>☐ National Professional Bodies<br>☐ NHS<br>☐ NICE<br>☐ Other (a box will open) |
| Please specify the other sources of information | __________________________________ |
| 3.10 Have you or any of your rehabilitation team had suspected or confirmed COVID-19 | ◯ Yes ◯ No<br>(Boxes will open for details) |
| Approximately how many staff | __________________________________ |
| What impact has this had on your service | __________________________________ |
| 4. REHABILITATION TEAM REFERRALS AND CASELOAD DURING THE COVID-19 PANDEMIC | |
| 4.1 Have your referral procedures or referral criteria changed during the COVID-19 pandemic | ◯ Yes ◯ No |
| Please give details | __________________________________ |
| 4.2 Has there been any change in the number of referrals to your service during the COVID-19 pandemic | ◯ A lot more<br>◯ Slightly more<br>◯ About the same<br>◯ A little less<br>◯ A lot less |
| Please give details on changes to the number of referrals | __________________________________ |
| 4.3 Has there been any change in the number of patients being seen by your service (your caseload) during the COVID-19 pandemic | ◯ A lot more<br>◯ Slightly more<br>◯ About the same<br>◯ A little less<br>◯ A lot less |
| Please give details on changes to the number of referrals | __________________________________ |
| 4.4 Would you say your face to face contact with patients is in general | ◯ A lot more<br>◯ Slightly more<br>◯ About the same<br>◯ A little less<br>◯ A lot less |
| 4.5 Would you say your telephone / remote connection contact with patients is in general | ◯ A lot more<br>◯ Slightly more<br>◯ About the same<br>◯ A little less<br>◯ A lot less |

| | |
|---|---|
| 4.6 Would you say your face to face contact with family members is in general | ◯ A lot more<br>◯ Slightly more<br>◯ About the same<br>◯ A little less<br>◯ A lot less |
| 4.7 Would you say your telephone / remote connection contact with family members is in general | ◯ A lot more<br>◯ Slightly more<br>◯ About the same<br>◯ A little less<br>◯ A lot less |
| 4.8 Would you say your contact with other members of the multi-disciplinary team is | ◯ A lot more<br>◯ Slightly more<br>◯ About the same<br>◯ A little less<br>◯ A lot less |
| 4.9 Would you say your telephone/remote connection advice and support for other clinicians is | ◯ A lot more<br>◯ Slightly more<br>◯ About the same<br>◯ A little less<br>◯ A lot less |
| 4.10 Have you furloughed any of your AHP team | ◯ Yes ◯ No<br>(A box will open for details) |
| Please give details | ______________________________ |
| 4.11 Have you lost staff from your service who have been redeployed to other services in your organisation or other organisations | ◯ Yes ◯ No<br>(A box will open for details) |
| Please give details (lost staff) | ______________________________ |
| 4.12 Have rehabilitation staff offered to help your service from elsewhere | ◯ Yes ◯ No<br>(A box will open for details) |
| Please give details | ______________________________ |
| 4.13 Have you changed how your rehabilitation service is organized (e.g. supporting patients with and without COVID-19) | ◯ Yes ◯ No<br>(A box will open for details) |
| Please give details | ______________________________ |
| 4.14 Have you changed where and how your staff work (including hours and location) | ◯ Yes ◯ No<br>(A box will open for details) |
| Please give details | ______________________________ |

| | |
|---|---|
| 4.15 Have you changed where and how your volunteers work (including hours and location) | ◯ Yes ◯ No<br>(A box will open for details) |
| Please give details | __________________ |
| 4.16 Have there been any changes to the patient facing information or instructional resources (online and paper) your team members are using during the COVID-19 Pandemic (including new resources and resources no longer used) | __________________ |
| 4.17 Please give details of any patient facing resources you feel are lacking | __________________ |
| 4.18 Please give detail of any other changes in the working practices of your team that are not covered in the questions above | __________________ |
| Use of virtual technologies | |
| 4.19 Are you are using virtual technologies (e.g. zoom, skype, teams) with patients and families to provide rehabilitation interventions | ◯ Yes, always used virtual technologies<br>◯ Yes, new service provision since start of COVID-19 pandemic<br>◯ No |
| What have been the difficulties with virtual technologies | __________________ |
| What has worked well with virtual technologies | __________________ |
| 4.20 Please give details of any virtual technology equipment provided for therapy team members to enable you to provide services and who provided it | __________________ |
| 4.21 Please give details of any virtual technology equipment provided for patients to enable rehabilitation services and who provided it | __________________ |
| 4.22 If no virtual technologies are being used to support the provision of rehabilitation services provided by your team members, or to support communication with patient and the wider MDT, please specify why | ☐ No access to technologies<br>☐ No training in use of technologies<br>☐ Rehabilitation interventions provided by this service, cannot be provided virtually<br>☐ Other (please explain below)<br>(Check all that apply) |
| Please give additional details | __________________<br>(Use this space for comments on any part of 4.22) |

5. CHALLENGES AND INNOVATION IN RESPONSE TO COVID-19

We want to know more about the challenges you have faced, their impacts on your service and care and how you have responded to them, including what you have found to be successful innovations

| | |
|---|---|
| 5.1 Have you had problems accessing personal protective equipment | ◯ Yes ◯ No<br>(Additional questions will open) |
| Please specify what you have had a shortage of | __________________________ |
| What did you do about it | __________________________ |
| Has this been a problem in the last 7 days | ◯ Yes ◯ Sometimes<br>◯ No |
| 5.2 Please give details if any patients are finding it more difficult to access services because of changes made to provision during the COVID-19 Pandemic | __________________________________ |
| 5.3 Please give details if any patients are finding it easier to access services because of changes made to provision during the COVID-19 Pandemic | __________________________________ |
| 5.4 Have you had any difficulties providing rehabilitation equipment / assistive devices | ◯ Yes ◯ Sometimes<br>◯ No<br>(The following questions may change) |
| Please give details about these difficulties | __________________________ |
| What did you do about problems accessing equipment / assistive devices | __________________________ |
| Have you had problems accessing equipment/assistive devices in the last 7 days | ◯ Yes<br>◯ Sometimes<br>◯ No |
| 5.6 Have there been other effects on yourself and/or members of your rehabilitation team that you think we should know about | __________________________________ |
| 5.7 Please tell us about any challenges and how you overcame them | __________________________________ |
| 5.8 What do you foresee will be the biggest challenges for COVID-19 in your rehabilitation service over the next months | __________________________________ |
| 5.9 What would help you to overcome these | __________________________________ |

6. Now please tell us about any innovations within your rehabilitation service (that are not already described above). We are keen to learn what has worked best for you

6.1 Please tell us about any change in practice or innovation that you think has been most successful to your working and the provision of rehabilitation by your team

__________________________________

6.2 Why is this

__________________________________

6.3 What would you say were the most important things that made this possible

__________________________________

6.4 Please list any other important innovations / changes you have made

__________________________________

6.5 What innovations do you plan to keep, to integrate into your usual service provision (including anything described in sections above)

__________________________________

7. FINALLY

Please indicate if you would like / are willing to be contacted regarding any of the following (Tick all that apply)

- ☐ To receive copy of the early reports and our newsletters as the findings emerge
- ☐ For us to check any information with you
- ☐ To be acknowledged as responding to this questionnaire (listed along with other services) in the reports and any publications
- ☐ To participate in any subsequent surveys like this one in the future when you may have made more changes or had more experiences

If you wish us to use a different Name and Email for the above (instead of the ones already given) please specify here

__________________________________

Please specify how you would like your service to be named and acknowledged in reports if applicable

____________________________

You will be free to opt out of receiving updates at any time, your details will not be passed onto other organisations or used for anything other than with your explicit consent above. Your individual responses will remain confidential, they will be analysed pseudo anonymously by the research team, with your service identified only by a code number unless you explicitly ask us to do otherwise.

Thank you for your help at this difficult time

PLEASE NOW CLICK SUBMIT

You must click "Submit" or "Save and Return Later" to save any data you have entered
