## Supplementary File 2 for "Understanding the impact of the Covid-19 pandemic on delivery of rehabilitation in specialist palliative care services: An analysis of the CovPall-Rehab survey data"

### Supplementary file 2: Procedures for CovPall-Rehab survey

Services were identified and contacted through national gatekeeper palliative care, hospice and allied health care professional organisations (see methods and acknowledgements for details) and asked that their rehabilitation lead, or their nominee, complete an on-line survey available via a link. The email attached the participant information sheet with details of the study rationale, ethical approval, data protection and management, investigators and contacts for further information or concern. No incentives were offered for completion. Completion was taken to indicate informed consent. The CovPall-Rehab study was presented at relevant online meetings and discussed with gatekeeper organisations to inform the methods, questions and to raise awareness and engagement.

We developed and piloted a secure, password-protected web-based data entry portal (in the Research Electronic Data Capture (REDCap) at the study coordinating centre, King’s College London. Services could keep their identity hidden if they wished, but most chose to provide an email for contact. Data were anonymised before analysis.

The questionnaire was developed and piloted by the CovPall study team building on an earlier survey of Italian hospices, adding questions on the impact of and response to COVID-19. It was intended to be brief, taking around 30 minutes to complete as we recognised staff were busy. The original CovPall survey was amended by the CovPall study team to focus on areas relevant to palliative rehabilitation. The amendments were approved by the Research Ethics Committee and comprised six sections: region and responder; palliative rehabilitation services offered pre-Covid-19; changes to palliative rehabilitation services during the pandemic, rehabilitation team referrals, caseload and changes to working practices during the pandemic; challenges and innovation for the provision of palliative rehabilitation services in response to Covid-19. Free-text explanatory comments were invited in all sections and respondents also indicated whether and how they could be contacted. Respondents could save and complete the questionnaire later if they wished. The questionnaire is available in supplementary file 1.

The survey opened on 30$^{th}$ July 2020 and closed on the 21$^{st}$ September 2020. The study coordinating centre replied actively to respondents’ queries or requests for help.

Data collection and entry was done electronically via the REDCap link directly by the Palliative Rehabilitation service leads without problems. No palliative rehabilitation service leads reported having difficulty accessing the online CovPall-Rehab survey and so did not require to take up the opportunity to complete the survey in paper format or over the telephone (with members of the research team at King’s entering the data). The research team audited the data weekly to ensure data entry completeness and sent monthly missing data and incomplete entry reports to the research associates and administrators, and where consent permitted to relevant respondents to check validity.
