## Supplementary File 3 for "Understanding the impact of the Covid-19 pandemic on delivery of rehabilitation in specialist palliative care services: An analysis of the CovPall-Rehab survey data"

**STROBE Statement—Checklist of items that should be included in reports of *cross-sectional studies***

| | Item No | Recommendation | |
|---|---|---|---|
| **Title and abstract** | 1 | (*a*) Indicate the study's design with a commonly used term in the title or the abstract | Yes |
| | | (*b*) Provide in the abstract an informative and balanced summary of what was done and what was found | Yes |
| **Introduction** | | | |
| Background/rationale | 2 | Explain the scientific background and rationale for the investigation being reported | Yes |
| Objectives | 3 | State specific objectives, including any prespecified hypotheses | Yes |
| **Methods** | | | |
| Study design | 4 | Present key elements of study design early in the paper | Yes |
| Setting | 5 | Describe the setting, locations, and relevant dates, including periods of recruitment, exposure, follow-up, and data collection | Survey opened on the 30/07/20 and closed on the 21/09/2020 |
| Participants | 6 | (*a*) Give the eligibility criteria, and the sources and methods of selection of participants | Yes |
| Variables | 7 | Clearly define all outcomes, exposures, predictors, potential confounders, and effect modifiers. Give diagnostic criteria, if applicable | Yes |
| Data sources/ measurement | 8* | For each variable of interest, give sources of data and details of methods of assessment (measurement). Describe comparability of assessment methods if there is more than one group | Yes, textual and quantitative variables described |
| Bias | 9 | Describe any efforts to address potential sources of bias | Yes – Page 5 Data were anonymised before analysis |
| Study size | 10 | Explain how the study size was arrived at | Yes – page 5 |
| Quantitative variables | 11 | Explain how quantitative variables were handled in the analyses. If applicable, describe which groupings were chosen and why | We used descriptive summary statistics (SPSS vs 24) presented in table 2. Page 7) |
| Statistical methods | 12 | (*a*) Describe all statistical methods, including those used to control for confounding | Not applicable |
| | | (*b*) Describe any methods used to examine subgroups and interactions | Not applicable |

| | | | |
|---|---|---|---|
| | | (*c*) Explain how missing data were addressed | Not applicable |
| | | (*d*) If applicable, describe analytical methods taking account of sampling strategy | Not applicable |
| | | (*e*) Describe any sensitivity analyses | Not applicable |
| **Results** | | | |
| Participants | 13* | (a) Report numbers of individuals at each stage of study—eg numbers potentially eligible, examined for eligibility, confirmed eligible, included in the study, completing follow-up, and analysed | The numbers potentially eligible could not be determined because this was an online survey. However, a total of 61 questionnaires were completed. |
| | | (b) Give reasons for non-participation at each stage | N/A |
| | | (c) Consider use of a flow diagram | N/A |
| Descriptive data | 14* | (a) Give characteristics of study participants (eg demographic, clinical, social) and information on exposures and potential confounders | Yes – see table 2 |
| | | (b) Indicate number of participants with missing data for each variable of interest | Not applicable |
| Outcome data | 15* | Report numbers of outcome events or summary measures | Yes – see table 2 |
| Main results | 16 | (*a*) Give unadjusted estimates and, if applicable, confounder-adjusted estimates and their precision (eg, 95% confidence interval). Make clear which confounders were adjusted for and why they were included | Not applicable. |
| | | (*b*) Report category boundaries when continuous variables were categorized | N/A |
| | | (*c*) If relevant, consider translating estimates of relative risk into absolute risk for a meaningful time period | N/A |
| Other analyses | 17 | Report other analyses done—eg analyses of subgroups and interactions, and sensitivity analyses | Yes<br>Page 9 -16 (Reflexive thematic analysis of free text responses) |
| **Discussion** | | | |
| Key results | 18 | Summarise key results with reference to study objectives | Yes<br>Pages 16-19 |
| Limitations | 19 | Discuss limitations of the study, taking into account sources of potential bias or imprecision. Discuss both direction and magnitude of any potential bias | Yes<br>Page 19 |
| Interpretation | 20 | Give a cautious overall interpretation of results considering objectives, | Yes page 19 |

| | | | |
|---|---|---|---|
| | | limitations, multiplicity of analyses, results from similar studies, and other relevant evidence | |
| Generalisability | 21 | Discuss the generalisability (external validity) of the study results | Yes |
| **Other information** | | | |
| Funding | 22 | Give the source of funding and the role of the funders for the present study and, if applicable, for the original study on which the present article is based | Yes – page 20 |

*Give information separately for exposed and unexposed groups.

**Note:** An Explanation and Elaboration article discusses each checklist item and gives methodological background and published examples of transparent reporting. The STROBE checklist is best used in conjunction with this article (freely available on the Web sites of PLoS Medicine at http://www.plosmedicine.org/, Annals of Internal Medicine at http://www.annals.org/, and Epidemiology at http://www.epidem.com/). Information on the STROBE Initiative is available at www.strobe-statement.org.

### Checklist for Reporting Results of Internet E-Surveys (CHERRIES)

| | **Checklist for Reporting Results of Internet E-Surveys (CHERRIES)** | | |
|---|---|---|---|
| ***Item Category*** | ***Checklist Item*** | ***Explanation*** | ***Page No.*** |
| **Design** | | | |
| | Describe survey design | Describe target population, sample frame. Is the sample a convenience sample? (In "open" surveys this is most likely.) | 5 Supplementary file 2 |
| **IRB (Institutional Review Board) approval and informed consent process** | | | |
| | IRB approval | Mention whether the study has been approved by an IRB. | 5 |
| | Informed consent | Describe the informed consent process. Where were the participants told the length of time of the survey, which data were stored and where and for how long, who the investigator was, and the purpose of the study? | 5 and Supplementary material file 2 |
| | Data protection | If any personal information was collected or stored, describe what mechanisms were used to protect unauthorized access. | 5, Supplementary file 2 |
| **Development and pre-testing** | | | |
| | Development and testing | State how the survey was developed, | 5, |

| | **Checklist for Reporting Results of Internet E-Surveys (CHERRIES)** | | |
|---|---|---|---|
| ***Item Category*** | ***Checklist Item*** | ***Explanation*** | ***Page No.*** |
| | | including whether the usability and technical functionality of the electronic questionnaire had been tested before fielding the questionnaire. | Supplementary file 2 |
| **Recruitment process and description of the sample having access to the questionnaire** | | | |
| | Open survey versus closed survey | An “open survey” is a survey open for each visitor of a site, while a closed survey is only open to a sample which the investigator knows (password-protected survey). | 5, Supplementary file 2 |
| | Contact mode | Indicate whether or not the initial contact with the potential participants was made on the Internet. (Investigators may also send out questionnaires by mail and allow for Web-based data entry.) | 5, Supplementary file 2 |
| | Advertising the survey | How/where was the survey announced or advertised? Some examples are offline media (newspapers), or online (mailing lists – If yes, which ones?) or banner ads (Where were these banner ads posted and what did they look like?). It is important to know the wording of the announcement as it will heavily influence who chooses to participate. Ideally the survey announcement should be published as an appendix. | 5, Supplementary file 2 |
| **Survey administration** | | | |
| | Web/E-mail | State the type of e-survey (eg, one posted on a Web site, or one sent out through e-mail). If it is an e-mail survey, were the responses entered manually into a database, or was there an automatic method for capturing responses? | 5, Supplementary file 2 |
| | Context | Describe the Web site (for mailing list/newsgroup) in which the survey was posted. What is the Web site about, who is visiting it, what are visitors normally looking for? Discuss to what degree the content of the Web site could pre-select the sample or influence the results. For example, a survey about vaccination on a | Supplementary File 1 and 2:<br><br>CovPall study webpages hosted on Cicely Saunders |

| | **Checklist for Reporting Results of Internet E-Surveys (CHERRIES)** | | |
|---|---|---|---|
| ***Item Category*** | ***Checklist Item*** | ***Explanation*** | ***Page No.*** |
| | | anti-immunization Web site will have different results from a Web survey conducted on a government Web site | Institute webpages on King's College London website. |
| | Mandatory/voluntary | Was it a mandatory survey to be filled in by every visitor who wanted to enter the Web site, or was it a voluntary survey? | Voluntary |
| | Incentives | Were any incentives offered (eg, monetary, prizes, or non-monetary incentives such as an offer to provide the survey results)? | No incentives were offered |
| | Time/Date | In what timeframe were the data collected? | 5 |
| | Randomization of items or questionnaires | To prevent biases items can be randomized or alternated. | N/A |
| | Adaptive questioning | Use adaptive questioning (certain items, or only conditionally displayed based on responses to other items) to reduce number and complexity of the questions. | Yes |
| | Number of Items | What was the number of questionnaire items per page? The number of items is an important factor for the completion rate. | Varied 6-10 depending on responses |
| | Number of screens (pages) | Over how many pages was the questionnaire distributed? The number of items is an important factor for the completion rate. | 12 |
| | Completeness check | It is technically possible to do consistency or completeness checks before the questionnaire is submitted. Was this done, and if "yes", how (usually JAVAScript)? An alternative is to check for completeness after the questionnaire has been submitted (and highlight mandatory items). If this has been done, it should be reported. All items should provide a non-response option such as "not applicable" or "rather not say", and selection of one response option should be enforced. | Yes, completeness checks were possible, and we were able to prompt respondents to give them the opportunity to complete the survey. |
| | Review step | State whether respondents were able to review and change their answers (eg, through a Back button or a Review step which displays a summary of the responses and asks the respondents if they are correct). | Yes. Respondents were given a unique access code and were able to save and return to the survey. |

| | **Checklist for Reporting Results of Internet E-Surveys (CHERRIES)** | | |
|---|---|---|---|
| ***Item Category*** | ***Checklist Item*** | ***Explanation*** | ***Page No.*** |
| | | | Supplementary files 2. |
| **Response rates** | | | |
| | Unique site visitor | If you provide view rates or participation rates, you need to define how you determined a unique visitor. There are different techniques available, based on IP addresses or cookies or both. | n/a |
| | View rate (Ratio of unique survey visitors/unique site visitors) | Requires counting unique visitors to the first page of the survey, divided by the number of unique site visitors (not page views!). It is not unusual to have view rates of less than 0.1 % if the survey is voluntary. | n/a |
| | Participation rate (Ratio of unique visitors who agreed to participate/unique first survey page visitors) | Count the unique number of people who filled in the first survey page (or agreed to participate, for example by checking a checkbox), divided by visitors who visit the first page of the survey (or the informed consents page, if present). This can also be called “recruitment” rate. | n/a |
| | Completion rate (Ratio of users who finished the survey/users who agreed to participate) | The number of people submitting the last questionnaire page, divided by the number of people who agreed to participate (or submitted the first survey page). This is only relevant if there is a separate “informed consent” page or if the survey goes over several pages. This is a measure for attrition. Note that “completion” can involve leaving questionnaire items blank. This is not a measure for how completely questionnaires were filled in. (If you need a measure for this, use the word “completeness rate”.) | n/a |
| **Preventing multiple entries from the same individual** | | | |
| | Cookies used | Indicate whether cookies were used to assign a unique user identifier to each client computer. If so, mention the page on which the cookie was set and read, and how long the cookie was valid. Were duplicate entries avoided by preventing users access to the survey twice; or were duplicate database entries having the same user ID eliminated before analysis? In the latter case, which entries were kept for analysis (eg, the first | n/a |

| | **Checklist for Reporting Results of Internet E-Surveys (CHERRIES)** | | |
|---|---|---|---|
| ***Item Category*** | ***Checklist Item*** | ***Explanation*** | ***Page No.*** |
| | | entry or the most recent)? | |
| | IP check | Indicate whether the IP address of the client computer was used to identify potential duplicate entries from the same user. If so, mention the period of time for which no two entries from the same IP address were allowed (eg, 24 hours). Were duplicate entries avoided by preventing users with the same IP address access to the survey twice; or were duplicate database entries having the same IP address within a given period of time eliminated before analysis? If the latter, which entries were kept for analysis (eg, the first entry or the most recent)? | n/a |
| | Log file analysis | Indicate whether other techniques to analyze the log file for identification of multiple entries were used. If so, please describe. | Identification for multiple entries was conducted by examining completed time stamped survey respondent details. No duplicate completed surveys were identified. |
| | Registration | In “closed” (non-open) surveys, users need to login first and it is easier to prevent duplicate entries from the same user. Describe how this was done. For example, was the survey never displayed a second time once the user had filled it in, or was the username stored together with the survey results and later eliminated? If the latter, which entries were kept for analysis (eg, the first entry or the most recent)? | Respondents logged in and were given a unique ID and password to return to the same survey. Respondents were able to get re login access details from research team if they forgot their survey log in details. |
| **Analysis** | | | |
| | Handling of incomplete questionnaires | Were only completed questionnaires analyzed? Were questionnaires which terminated early (where, for example, users did not go through all questionnaire pages) | Only submitted timestamped surveys were |

| | | | |
|---|---|---|---|
| | **Checklist for Reporting Results of Internet E-Surveys (CHERRIES)** | | |
| ***Item Category*** | ***Checklist Item*** | ***Explanation*** | ***Page No.*** |
| | | also analyzed? | analysed. Surveys with missing data items were included in the analysis. |
| | Questionnaires submitted with an atypical timestamp | Some investigators may measure the time people needed to fill in a questionnaire and exclude questionnaires that were submitted too soon. Specify the timeframe that was used as a cut-off point, and describe how this point was determined. | N/A |
| | Statistical correction | Indicate whether any methods such as weighting of items or propensity scores have been used to adjust for the non-representative sample; if so, please describe the methods. | No statistical corrections were conducted. |
