## Supplementary File 4 for "Understanding the impact of the Covid-19 pandemic on delivery of rehabilitation in specialist palliative care services: An analysis of the CovPall-Rehab survey data"

*Supplementary Material 2: Further Illustrative Quotes by theme*

| Themes | Illustrative Quotes |
|---|---|
| **Theme 1: Fluctuating shared spaces.** | *"Day Hospice closed so all face-to-face interventions with patients in the community immediately stopped. Initially not allowed to go into community to see patients" (ID08 England*<br><br>*"Patients not able to have assessments/interventions in the hospice as closed, carried out virtually in their homes instead" ID51 England)*<br><br>*"No face to face input with existing patients and no new patient referrals taken for rehab" (ID17 England)*<br><br>*"No major changes – both our Occupational Therapists have continued to support people in their own homes during the pandemic." (ID03 England)*<br><br>*"Still sees patients on the in-patient unit and in home setting if required. Still see individuals with urgent requirements such as a home assessment prior to discharge". (ID02 England)*<br><br>*"No change – usual rehabilitation and enablement approach" (ID28 England)* |
| **Theme 2: Remote and digitised rehabilitation offer** | *"use of AccRx on SystmOne* [clinical online virtual platforms] *for video consultations, sending out more postal information to patients. Zoom recorded and live groups sessions". (ID09 England)*<br><br>*"Increased the use of video recordings shared on Facebook and sent to patients. Physio exercises included in patient newsletters. Created YouTube education talks" (ID29, England)*<br><br>*"Day Hospice & breathlessness service patients; some assessments of patients in own homes completed/ commenced via video calls. Visits into patients' homes only where absolutely necessary".*<br><br>*"Videos produced to support people in self-help activities, exercises and practical advice. Handouts produced to send to those who do not wish to / cannot access the internet." (ID02, England)*<br><br>*"For all out patient /community appointments provided virtually by telephone or AccuRx unless clear clinical need to visit F2F. Information about breathlessness management provided on website and group sessions provided for day therapy offered as part of virtual day therapy" (ID39)* |

| | |
|---|---|
| **Theme 3: Capacity to provide and participate in rehabilitation** | **Sub-theme 1: Reach and access**<br>*“As the hospice is in an awkward place to get to geographically - virtual has enable people who did not want to travel to participate. Some people have enjoyed the Zoom classes especially when they would have not been well enough to travel - they would sometimes miss outpatient and day therapy appointments as the journey was too long” (ID13, England)*<br>*“Patients who have been wary of coming to the hospice have found it a far more acceptable way to be introduced to our services/ teams. The use of Zoom has been a benefit in assessing patients progress and being able to refer for further support where appropriate (as opposed to phone assessment). One patient with MND has benefitted greatly from the opportunity to engage in group sessions and communication challenges have been accommodated very well by the group” (ID14, England)*<br>Younger patients with better access to online services have found benefit from not being able to access classes online rather than coming out to appointments. We restarted our community provision (physio) in order to continue to provide a face to face service enabling access for some people who may otherwise have come to an outpatient appointment. *(ID07)*<br>*“not all patients have the necessary IT to access change of services, or some prefer not to use this access route - so need to offer either face to face or telephone support.” (ID09 England)*<br>*“People who don't want to engage with technology some people only want to see someone in person” (ID29, England)*<br>*“Some patients are not tech savvy or do not have access to internet so in early weeks service to them was limited to phonecalls unless in emergency situations. Huge impact on Day Centre patients, especially mental wellbeing for those that live alone” (ID12 England)*<br>*“Physiotherapy and Occupational Therapy have access to use Zoom for patient assessments however have found that it is not useful for the majority of our client group” (ID05 England)*<br>**Sub-theme 2: Rapid redeployment and disrupted resources**<br>*“Redeployment to HCA role - able to train healthcare assistants on the job in rehab and enablement approach” (ID28 England)*<br>*“Our band 4 physio tech was initially redeployed onto the ward as a nursing assistant and also to provide some physio cover. She is now helping me to run virtual rehab classes to our patients in* |

| | |
|---|---|
| | *the community. Our band 3 rehab assistant initially was redeployed onto the ward but is now back in her usual role assisting the Occupational Therapists (ID03 England)*<br><br>*"Redeployment, followed by completely redesigning service and its delivery has left the team exhausted but still committed to patient care" (ID16 England)*<br><br>**Sub-theme 3: Emotional and physical distress**<br><br>*"Responding to constant changes initially was challenging to support the team with. All team members have had to adapt to some degree how they perform their role. We had an office move to allow social distancing". (ID45, England)*<br><br>*"difficult for staff to manage disappointment form patients that services are not able to be provided 'as usual'. (ID38, England)*<br><br>*"Having been used to being very active the sedentary nature of providing therapy via a screen has been exhausting. Headaches and muscular aches have been common. Having to wear PPE is exhausting particularly having to overcome the challenge of communicating via a mask" (ID14, England)*<br><br>*"It has been emotionally taxing for the team to manage patient expectations or referrals that may not be normally appropriate for our services but with the knowledge other services cant take the cases on they have worked above and beyond to make sure patients are supported as best as they can. The closure of day services has had a lasting impact" (ID52, England)*<br><br>*"increased colleague conversations to enable right professional at right time visits has been beneficial.... arranged team Covid debrief was very beneficial in last 2 weeks" (ID25, England)* |
| **Theme 4: Covid-19 as a springboard for positive change** | *"videoconference access to individual and group therapy sessions will be integrated into our service in the future. We have also increased our confidence in the patient reported assessments - we intend to continue empowering patients when possible to do this." (ID14, England)*<br><br>*We are looking into the possibility of ongoing provision of seven day therapy services … We are working towards the Community Teams being more integrated, with therapists embedded in the teams. (ID38, England)*<br><br>*"looking about how we could use our Compassionate Neighbours to support rehab in the community thinking about whether a green gym may be possible in local park area how to upskill more volunteers to support" (ID01, England)* |

| | |
|---|---|
|  | *"It is likely that online exercise group will continue for the foreseeable future, as will online Day Therapy due to covid restrictions. Patients however want face to face contact and it may not last long term due to difficulties engaging new referrals into the online group". (ID05 England)*<br><br>*"We used to fully discharge individuals but they could re-refer themselves - we have recognised that some individuals benefit from regular check-ins and for those where this is warranted we would continue. We have recognised that for many the impact of telephone input can be sufficient, meeting their needs and proving very convenient. Video assessments and input also more convenient for some so could continue so long as cost of things like accuRx which is currently free do not become restrictive". (ID30 England)*<br><br>*"Definately virtual classes and we hope to increase these over the next few weeks. Also the care home support will continue as the feedback we got was excellent". (ID03)*<br><br>*"more thorough triage processes and phone assessments to establish clear need and purpose of any visits". (ID25)* |
